# Anthelminthic treatment modifies cytokine profiles and improves the performance of tuberculin test in helminth-*Mycobacterium bovis* co-infection in cattle herds

**DOI:** 10.1101/2024.04.02.24305188

**Authors:** Victor O. Akinseye, Theophilus A Jarikre, Moshood O Tijani, Olugbenga O Alaka, Simeon I. Cadmus

## Abstract

While enormous efforts are directed at tackling the problems of human tuberculosis (TB) globally, the aspect of bovine TB (bTB) has received limited attention particularly in developing countries, where control policies are non-existent or inadequate. We compared the results of a single intradermal comparative cervical tuberculin test (SICCTT) with cytokine profiles before and after anti-helminthic treatment in cattle in their natural environment. Cattle were initially subjected to SICCTT and their helminth status assessed by coprological testing for *Strongyle-type, Strongyloides sp*., *Toxocara vitulorum, Nematodirus sp*., *Paraphistomum cervi, Fasciola gigantica, Dicrocoelium dendriticum and Moniezia benedeni*.; 60 days after, they were dewormed and the SICCTT was repeated with coprological testing; SICCTT-positives underwent post mortem examination and mycobacterial culture to confirm TB infection. Also, we measured the level of circulating type 1 (gamma interferon [IFN-γ]), type 2 (interleukine-4 [IL-4]) and regulatory (transforming growth factor-beta [TGF-β]) cytokines before and after anthelminthic therapy. Results revealed that cattle positive by SICCTT showed typical granulomatous lesions suggestive of TB which was confirmed by positive culture. The proportion of SICCTT positive cattle increased to about two folds following anthelminthic treatment. Cattle co-infected with helminth-bTB have significantly (p = 0.000) lower levels of IFN-γ and higher levels of IL-4 and TGF-β. Conversely, anthelminthic therapy led to significantly (p=0.000) elevated levels of IFN-γ, concomitant (p =0.000) decreased levels of IL-4 and TGF-β. Overall, we show that anthelminthic treatment can improve the rate of SICCTT positivity and reverse the modulation of cytokine responses in helminth-bTB co-infection.

## INTRODUCTION

Bovine tuberculosis (bTB) is an important disease of cattle caused by *M. bovis*. The disease is more prevalent in low- and middle-income countries, where it shares the same geographical niche with helminth infection. The existence of multiple co-infections is an expected occurrence in natural populations, where an animal is infected with multiple pathogens (1). Several parasites are known to have the capability of modulating the host immune response to further their own existence (2). Notably, this results in the alteration of the host response to co-infecting pathogens, with consequent far-ranging effects on the transmission and progression of the disease as well as diagnostic test accuracy (2, 3). However, the outcome of these interactions is dependent on combinations of variable factors which are peculiar to individual host (4). *Mycobacterium* infection, like many other infectious diseases, has hitherto been largely studied in the absence of co-infection. Nevertheless, co-infection with different classes of helminths has been identified as a contributor to the poor efficacy of bTB diagnostic tests performance (5). Helminths like *Fasciola hepatica*, a widespread trematode of livestock is known to induce anti-inflammatory, Th2 response in its host (5). This predominant Th2 response has been shown to down-regulate Th1 pro-inflammatory responses with resultant dampening of interferon-gamma (IFN-γ) production, a mechanism that is essential for the parasite survival within the host (6, 7, 8).

In many developed and high-income countries, like New Zealand and the United Kingdom, the “test and slaughter” bTB diagnostic programme has contributed positively in reducing the incidence of *M. bovis* infection in livestock populations (5). The diagnostic regimen uses the single intradermal comparative cervical tuberculin test (SICCTT) to identify infected animals; culling is then initiated in animals that test positive. Important antemortem diagnostic tests such as SICCTT and IFN-γ assays measure delayed-type hypersensitivity response to purified protein derivative (PPD)-a tuberculin antigen that is dependent on active Th1 cell and associated secretion of IFN-γ in response to *M. bovis* infection (9). Notably, no gold standard exists for the ante-mortem detection of bTB, with all available tests fraught with varied and relatively different degrees of sensitivity (10) which has been attributed to several factors (9, 11, 12, 13). However, a major issue that has been poorly investigated in livestock population is the presence of concurrent infection with pathogens that may impair the immune response to *M. bovis* infection and possibly affect SICCTT reactivity and IFN-γ production (4, 5, 9).

Helminths generally are common parasites of livestock worldwide, with the highest level of prevalence being observed in low- and middle-income countries. This is partly due to poor practices that facilitate the spread of infections among animal handlers such as communal land use which encourages common feeding paths and watering points (4). Furthermore, poor animal health care systems and lack of effective drugs coupled with the misuse of drugs by farmers or unqualified personnel are other factors attributed to the high incidence of helminths in this part of the world (14). Further, helminths are known to cause asymptomatic or subclinical protracted infections that usually elicit anti-inflammatory immune response characterized by unregulated Th2 and T regulatory cell (Treg cells) cytokine production that down-regulate Th1 cytokines response (14). Several authors have investigated the possible role of helminths on the infection outcome and diagnosis of TB in humans (15, 16, 17, 18) and experimental animals (5, 18, 19, 20). Many parasites can modulate the host immune response in order to further their own survival (1). This also alters the host response to co-infecting pathogens and can have wide-ranging effects, from the transmission and progression of the disease to the accuracy of diagnostic tests (2, 3).

While the majority of these studies believe that helminths impair the diagnosis of TB (5, 6, 7, 8), very few hold contrary view (21). Furthermore, not too long ago, the important role of regulatory T cells in the diagnosis and outcome of infection in TB were identified in humans (17). However, out of oversight, less priority or total neglect, few of the studies have considered livestock population. Again, the majority of the investigations conducted in the area of helminth-TB co-infection have been focused on the human populations, while only a few have involved cattle. The few studies on cattle were mainly in experimental settings, with only one study based on cattle in their natural conditions (22). Interestingly, none of these studies investigated the effect of anthelminthic treatment on the performance of SICCTT and the cytokines profile of cattle in their natural conditions and environmental settings.

In this study, we showed that anthelminthic treatment can enhance the diagnostic performance of the SICCTT in cattle within their natural environment in a developing country. Importantly, we report the down-regulation of TGF-β and IL-4 and up-regulated levels of IFN-γ, following anthelminthic treatment in co-infected cattle under a natural condition in a typical tropical African setting in Nigeria. There is, therefore, an urgent need for more studies to be carried out to elucidate the role of helminths in the accurate diagnosis of bTB among cattle under natural condition considering the ongoing global effort at tackling the scourge of TB in humans from the zoonotic transmission perspective (23).

## METHODS

### Study population and study design

The study population comprised of cattle from two private herds in a university community. Invariably, these cattle were originally sourced from Fulani herds. The Fulanis are itinerant pastoralist and agro-pastoralist who have migrated with their cattle herds from the northern region of the country in search of pastures and water, especially during the dry season, and have become domiciled in the south. All the cattle in the two herds, amounting to 54 in total were screened using the SICCTT. Blood and faecal samples were collected from individual cattle; the blood collected was used to quantify circulating cytokines (*ex-vivo*) and faecal samples were examined for helminth burden (Figure 1). The study was divided into two phases: the first phase involved the screening of cattle with the SICCTT and collection of blood and faecal samples. The second phase which was carried out 60 days after the first SICCTT, involved treatment of cattle with anti-helminthic (Albendazole [Albenol-600 bolus MBA2], Interchemie, The Netherlands; based on manufacturer’s instructions); and between one to two weeks later, another SICCTT, and repeat blood and faecal sample collections were carried out. Individual animals were considered as a positive reactor if “Bovine minus Avian tuberculin” reading was ≥4mm (B-A ≥4mm). The effect of albendazole was evaluated by repeated coprological testing of the faecal samples collected after treatment with anthelminthic.

**Figure 1.**
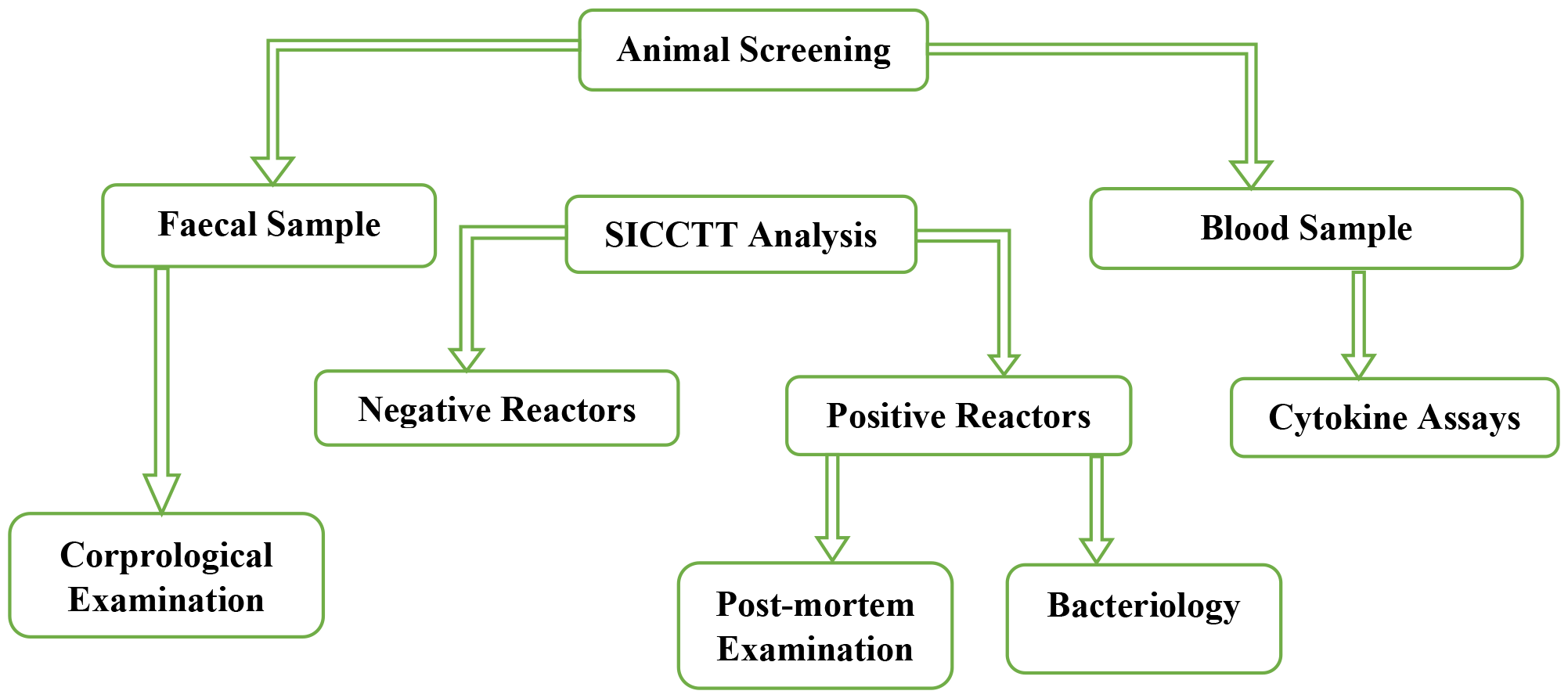
Experimental flowchart for the study

**Figure 2:**
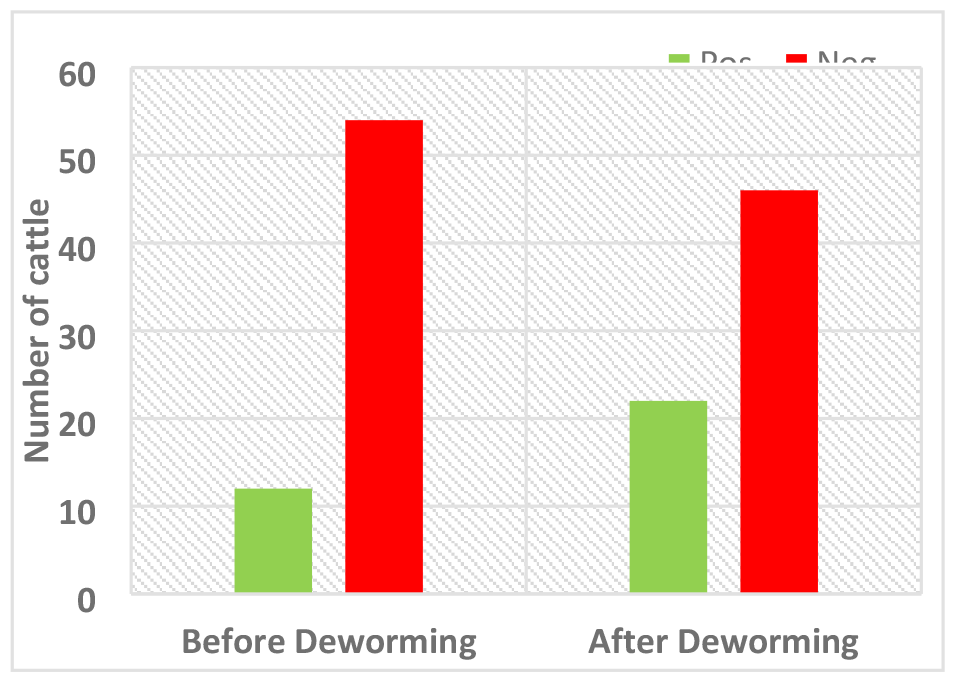
Total number of positive and negative reactor cattle before and after deworming.

### Diagnostic tests

#### Single intradermal comparative cervical tuberculin test (SICCTT)

Cattle were intradermally injected with both bovine and avian purified protein derivatives (PPDs) in the middle neck region (on the left side for consistency) as described by the World Organization for Animal Health standards (24). Briefly, two sites of about 2 cm^2^ in area, approximately 12 cm apart, were shaved using a sterile surgical blade after which the skin thickness was measured using a vernier calliper. Thereafter, exactly 0.1 mL containing 30,000 IU/mL of B-PPD (bovine tuberculin PPD, Prionics Lelystad BV, Lelystad, The Netherlands) and 0.1 mL with 25,000 IU/mL of A-PPD (avian tuberculin PPD, Prionics Lelystad BV, Lelystad, The Netherlands) were intradermally injected with the use of two separate sterile syringes (one for each) into the shaved area. At each injection site, the formation of a bleb confirmed a successful procedure. After 72 h, measurement of the shaved areas among animals screened was done using the vernier calliper. The initial and final measurements were carried out by the same person to avoid the error which may arise from individual measurement variations.

### Coprological examination

The faecal samples of cattle were collected in a sterile cellophane paper and transported to the laboratory under cold chain and stored at 4°C until analysed. Floatation and sedimentation tests were carried out on each sample to identify eggs (*Strongyle-type, Strongyloides sp*., *Toxocara vitulorum, Nematodirus sp*., *Paraphistomum cervi, Fasciola gigantica, Dicrocoelium dendriticum and Moniezia benedeni*) as well as copro-culture to identify larvae as previously described (25).

### Post-mortem Examination and Mycobacterial Culture

Detailed post-mortem examination was carried out according to the World Animal Health Organisation (OIE) procedures (24) on the SICCT positive cattle, and suspected tuberculous lesions were identified and obtained aseptically for further confirmatory analyses. Samples collected were placed in labelled sterile screw-capped, leak-proof specimen containers and frozen at -20^0^C until they were processed. The tissues, processed as earlier described (26, 27), were inoculated on Lowenstein-Jensen slopes with pyruvate and/or glycerol and incubated at 37ºC for 12 weeks. All cultures were examined daily for the first 7 days after incubation to detect rapidly growing mycobacteria (non-tuberculous mycobacteria) and also to detect contamination. Thereafter, the cultures were examined once a week for 8 to 12 weeks to detect positive cultures of mycobacteria before adjudging the culture to be negative if there was no growth. Colonies from all resultant growth (Figure 3A) were examined for morphological appearance and acid-fast properties.

**Figure 3:**
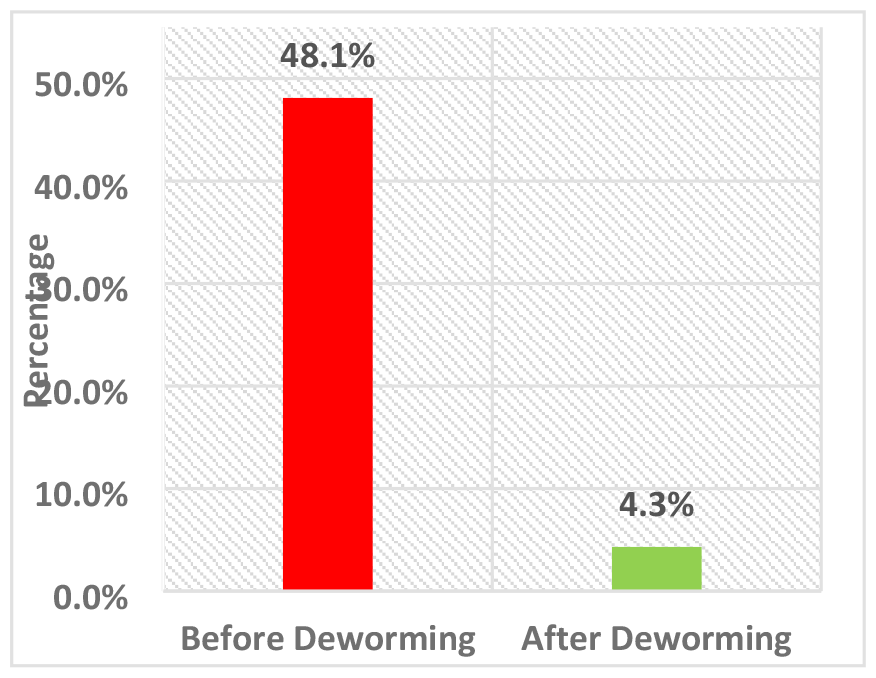
Proportion of helminth load before and after deworming.

### Cytokine Assays

#### Interferon-gamma (IFN-γ) assay and Interleukin 4 (IL-4)

Interferon-gamma (IFN-γ) and interleukin-4 (IL-4) levels in the serum obtained from whole blood of cattle were quantified with a standard ELISA technique using commercially available kits (Mabtech, Nacka Strand, Sweden) according to the manufacturer’s instruction. Briefly, high protein binding ELISA NUNC MAXISORP 96 well plates were coated with a monoclonal antibody (mAb) and incubated overnight at 4°C. On day 2, plates were washed twice with PBS and afterwards blocked with PBS+0.05%Tween 20 containing 1% BSA for one hour. After this, plates were washed five times with PBS containing Tween 20 and 100µl of neat serum samples and standard was added to all wells accordingly and incubated at room temperature for 2 hours. The plates were washed again five times and 100µl of biotinylated mAb was added to all wells and incubated for one hour at room temperature. After washing for five times, 100µl of Streptavidin-Alkaline phosphate conjugate was added to all wells and incubated at room temperature for one hour. The plates were washed again and 100µl of P-nitrophenyl-phosphate (pNPP) was added to all wells and the plates were read at 405nm by an ELISA reader after 30 minutes of developing time.

### Transforming growth factor-beta (TGF-β)

Transforming growth factor-beta (TGF-β) levels in the serum of cattle were quantified with a standard competitive ELISA technique using a commercially available kit (MyBioSource San Diego, California, USA) according to the manufacturer’s instructions. Briefly, 100µl of neat samples and standards were added to plates and PBS was used as blank control. Afterwards, 50µl of the conjugate solution was added to all wells except the blank control wells. The plates were then incubated for one hour at 37°C, after which they were washed five times. 50ul of substrate (substrate A and substrate B) solutions were added and incubated for 20 minutes at 37°C. 50µl of stop solution was added to all wells and plates were read with BioRad microplate reader at 450nm. All the optical density (O.D.) values were subtracted from the mean value of the blank control before result interpretation. The standard curve was constructed by plotting the concentration against the average of O.D. for each standard. The concentrations of samples were then calculated to correspond to the mean absorbance from the standard curve.

### Statistical Analysis

Data were analyzed using the STATA software version 12. The number of replicates, mean, standard deviation and standard error of mean were calculated. Group comparisons were statistically compared using paired t-test statistic, two-tailed, with statistical significance set at <0.05.

### Ethical consideration

Ethical permit for this study was given by the University of Ibadan-Animal Care and Use Research Ethics Committee (No. UI-ACUREC/19/138). Furthermore, all methods were performed following the relevant guidelines and regulations.

## RESULTS

### Response to SICCTT and anthelminthic treatment

The SICCTT was performed on all cattle, and sixty days later after anthelminthic treatment, the test was repeated. We found a significant increase in the number of positive reactors (P= 0.000) after the cattle were dewormed. The proportion of cattle that showed positive reactivity to SICCTT increased from 22.2% (12/54) to 47.8% (22/46) following anthelminthic treatment, giving a percentage increase of 45.5% (Figure 2). It was observed that seven (7) out of the 12 previously positive reactors (58%) showed an increase in the degree of positivity (PPDB minus PPDA response) after deworming. Further, significant decrease was observed in the worm load of animals screened after anthelminthic treatment (P=0.00). The worm load reduced from 48.1% to 4.3%, giving a percentage decrease of 90.9% (Figure 3).

In all, data was available for 46 cattle of the original 54 cattle, which were retrospectively divided into four groups based on SICCTT status and the detection of helminths. These include:

Group 1 (Control, n=8): these were cattle without bTB and helminth infection, but were subjected to anthelminthic treatment;

Group 2 (bTB infected cattle, n=12): these were cattle that were positive for mycobacterial infection based on tuberculin screened test;

Group 3 (helminth infected cattle, n=22): these were cattle that harboured at least one helminth egg or larvae;

Group 4 (bTB-helminth coinfection, n=4): these were cattle that were both positive to mycobacterial infection and helminth infection.

### Impact of anthelminthic treatment on the cytokine profiles of cattle

The results showed significant elevated concentration of circulating IFN-γ in cattle infected with bTB (group 2) compared to those with helminth infection (group 3), bTB-helminth coinfection (group 4) and the control (group 1) respectively (Figure 4). However, following anthelminthic treatment, there was an observed significant increase in the level of IFN-γ in co-infected cattle, whereas, no significant difference was seen in the circulating IFN-γ concentration of helminth infected and the control (Figure 4). Also, the concentration of IFN-γ increased in bTB infected cattle, but the difference was not significant (Figure 4).

**Figure 4:**
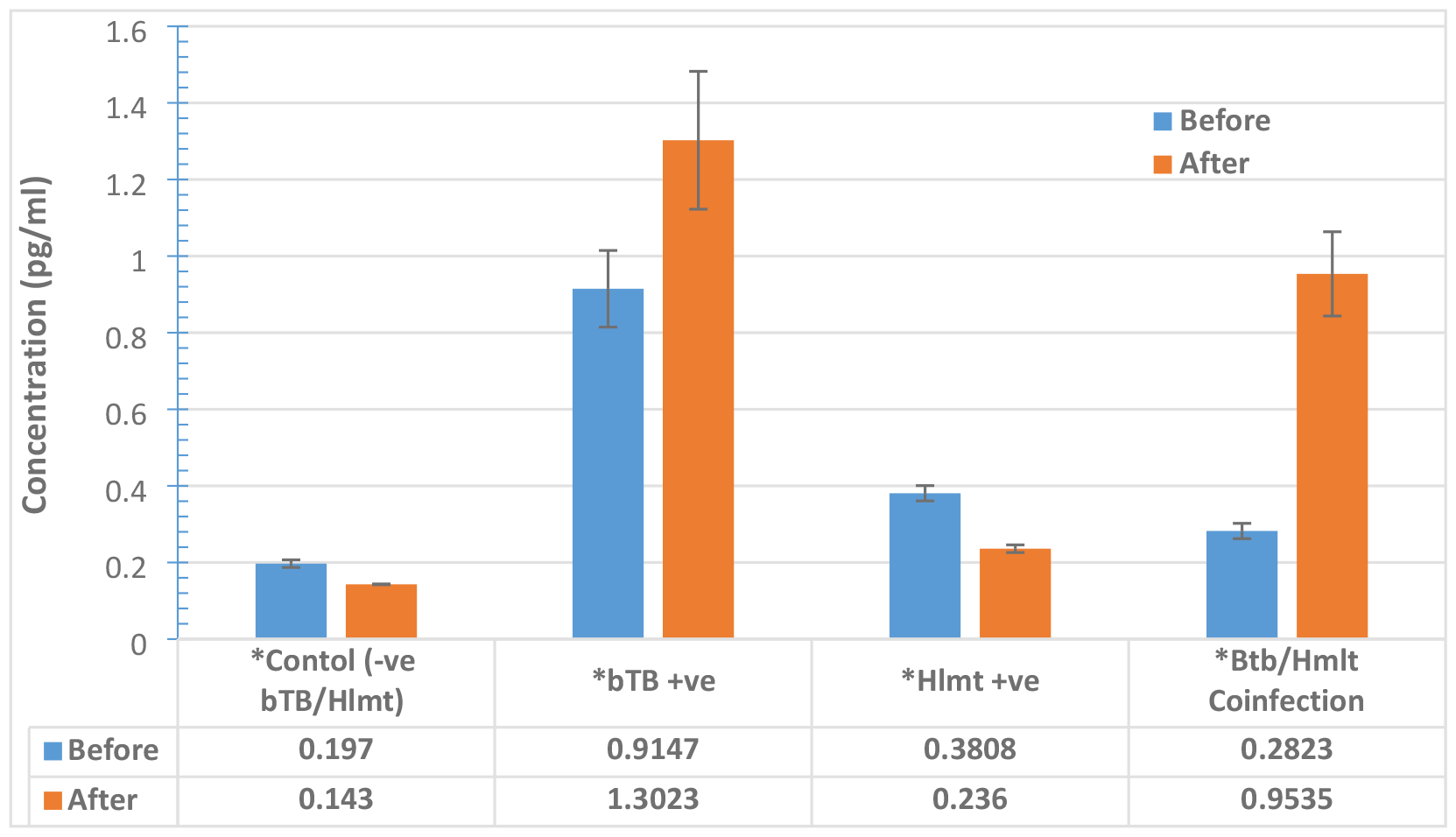
Total level of circulating IFN-γ of cattle screened before and after anthelminthic treatment, presented in Mean±S.D.

The results of the IL-4 and TGF-β followed the same trend. A significant elevated level of circulating IL-4 and TGF-β were observed in cattle having helminth infection (group 3) and those with bTB-helminth co-infection (group 4) compared to those with bTB infection (group 2) and the control (group 1) (Figure 5 & 6). However, following anthelminthic treatment, the concentration of circulating IL-4 and TGF-β significantly decreased and the reduction were comparable to those of the bTB infected group (group 2) and the control (group 1) (Figure 5 & 6). No significant difference was seen in the level of these two cytokine before and after anthelminthic treatment (Figure 5 & 6).

**Figure 5:**
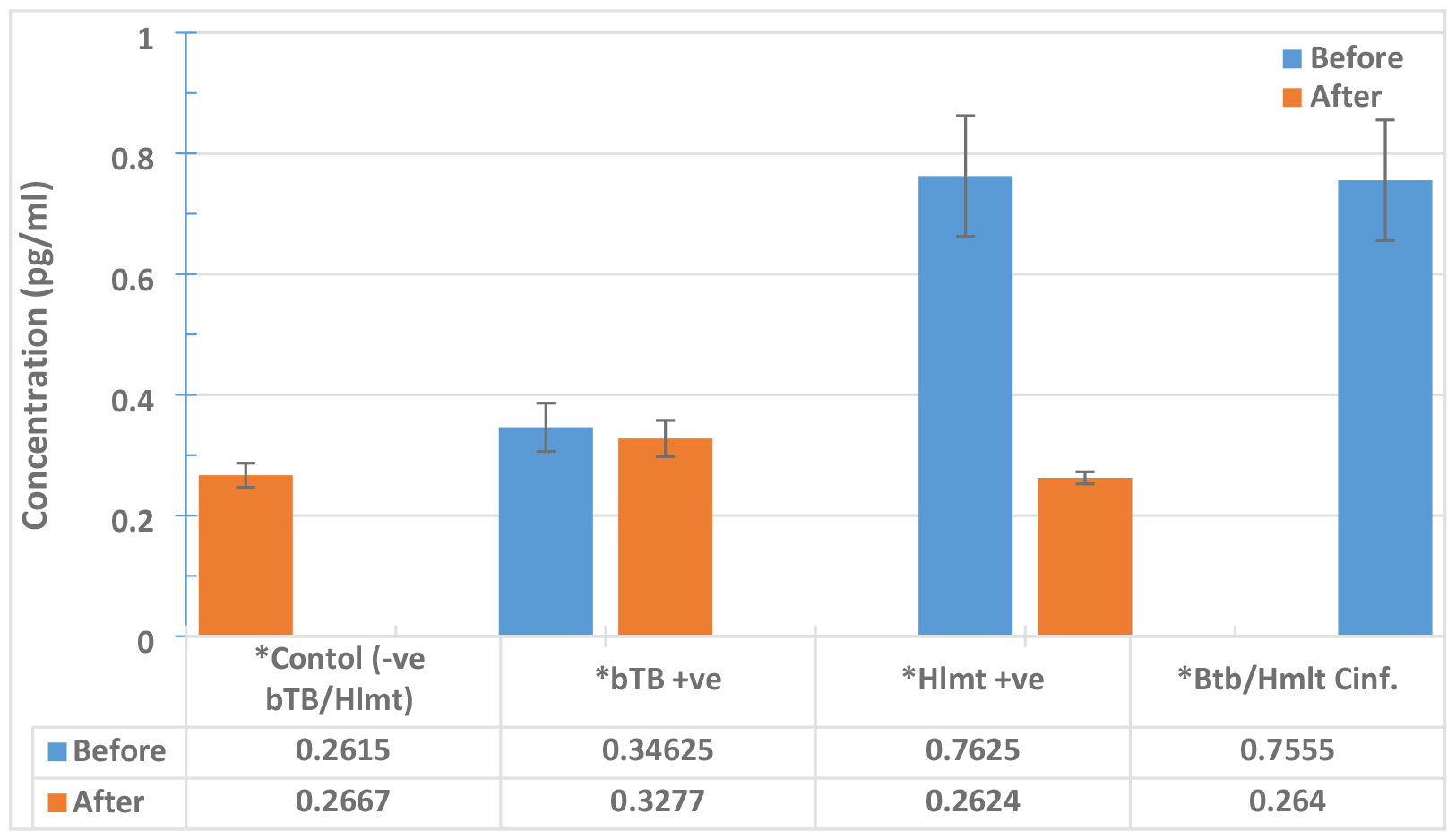
Total level of circulating IL-4 of cattle screened before and after anthelminthic treatment, presented in Mean±S.D.

**Figure 6:**
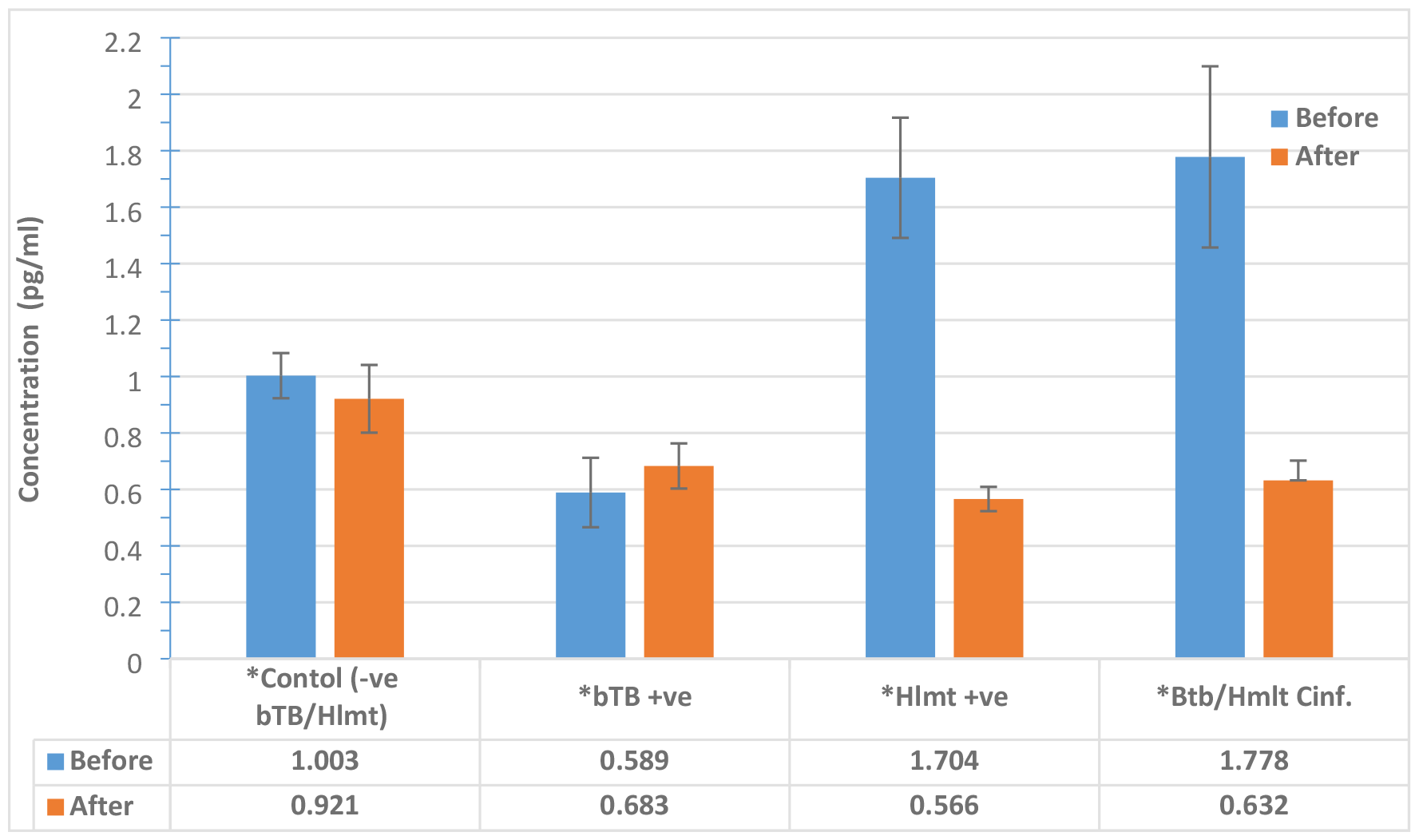
Total level of circulating TGF-β of cattle screened before and after anthelminthic treatment, presented in Mean±S.D.

### Post-mortem and culture

Following SICCTT positive reactivity in cattle (Figure 7A), post-mortem examination of SICCTT-positives (n=8) revealed an enlarged bronchial and mediastinal lymph nodes which were hyperaemic and firm with yellowish gritty nodules on the cortices of the lymph nodes. Also, their lungs were non-collapsed and consolidated with patchy prominent lobules having yellowish gritty nodules (1 to 5 cm diameter) within the parenchyma of the lungs (Figure 7B & 7C). Microscopically, the nodules were localised foci of chronic inflammatory reaction (granuloma) comprising centre of caseous necrosis surrounded by activated macrophages, lymphocytes, plasma cells, and delineated by fibroblasts with in the alveolar spaces. Acid-fast bacilli were observed within the granuloma, both in macrophages and extracellularly in the airspaces. The culture medium showed colonies typical of *Mycobacterium* growth and these were confirmed by acid stain (Figure 7D).

**Figure 7A.**
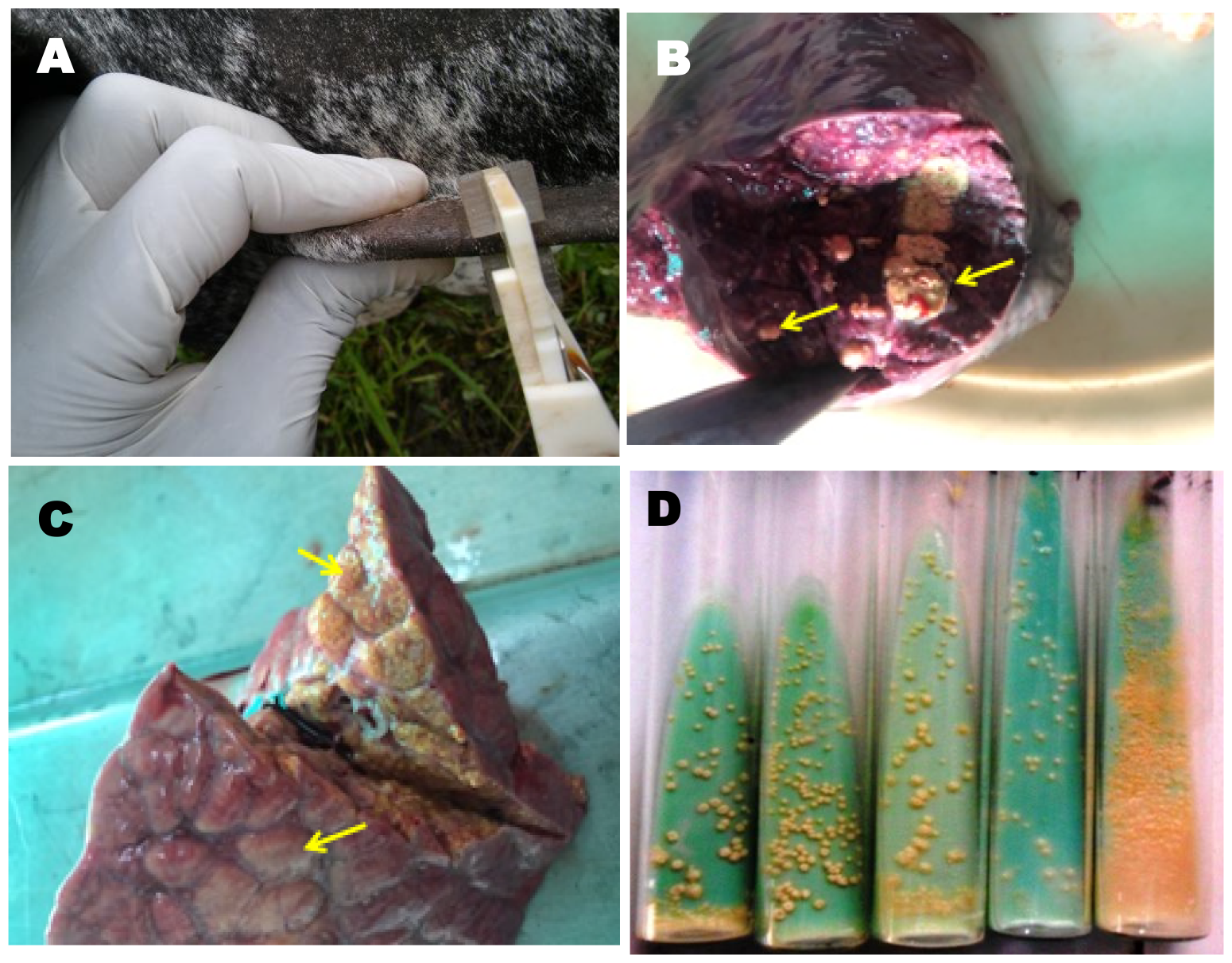
The SICCTT showing induration indicating a positive reaction. **Figure 7B& 7C: Yellowish, gritty nodules in the lung parenchyma from two different cattle indicative of TB.** **Figure 7D: A positive *Mycobacterium* growth**

## DISCUSSION

The immune response against *M. bovis* infection in cattle is typified by strong pro-inflammatory Th1 response and associated cytokines like IFN-y and IL-2 (28). Concurrent helminth infection can modulate the immune response by skewing it towards Th2 and/or regulatory T cell anti-inflammatory dominance (5). Previous studies in experimental animals have shown a negative association between helminthic and mycobacterial infections with an evident down-regulation of Th1 response and associated reduced IFN-γ production, lymphocyte responsiveness as well as up-regulation of the Th2 cytokines, IL-4 and IL-5 (5, 28, 29). These studies reiterate the fact that helminths generally tend to modulate the host immune response, to protect itself from destruction, and evade detection and eventual expulsion from the host (20, 30).

The findings of this small study revealed high prevalence of bTB among the cattle screened using SICCTT, supported by post-mortem and culture, the gold standard test for TB diagnosis. It was observed that anthelminthic treatment significantly increased the proportion of positive reactors to SICCTT suggesting enhancement of Th1 response (P=0.001). This is in agreement with a previous study where a significant improvement in the response to the tuberculin skin test in humans was reported following treatment with anti-helminthics (15). Similarly, improvement in T cell responses to PPD and IFN-γ production was observed in filarial worm-infected people after treatment with anti-helminthics (31, 32, 33). However, the findings of this study are in contrast to that of Byrne e*t al*, (2019) where no impact of liver fluke was found in animals being screened for bTB, using SICCTT, at an individual level, in a large scale retrospective surveillance dataset study (34). The difference in the conclusions arrived at may not be unconnected with the long average time between test and slaughter (i.e. 106.84 days) and non-consideration of valuable data on the administration of flukicide in the analyses used by Byrne *et al*. (2019) (34). Further, it is noteworthy, in the present study, that about 58% (7/12) of the previously SICCTT-positive cattle showed an increase in the degree of positivity after deworming. It is known that helminth infection can reduce weak SICCTT-positive reactors into being classified as SICCTT-negative in bTB-coinfected cattle (5). Results of this study clearly showed that anthelminthic treatment was not only able to reverse this trend but resulted in converting weak SICCTT positive animals to more positive reactors.

Significant increase in the level of circulating IFN-γ was seen following reduction or elimination of intestinal worms after treatment with the anthelminthic drug in co-infected cattle (group 4), and those with helminth infection (group 4). Importantly, the level of IFN-γ after anthelminthic treatment was comparable to that of cattle infected with only bTB (group 2). Significantly increased proliferation of Th1 cell and IFN-γ production following anthelminthic therapy have been reported in a previous clinical study (11) with associated skewed immune response dominance toward Th2 dominance prior to anthelminthic exposure. Experimental studies in cattle have been reported to show downregulation of Th1 antimicrobial response (mainly, IFN-γ) in co-infected cattle with associated up-regulation of the Th2 cytokines IL-4 and IL-5 (28, 35, 36). In the present study, increased concentration of IFN-γ observed after anthelminthic treatment could have resulted from the reduced dominance of Th2 and other helminth-driven immunomodulatory immune responses in co-infected cattle culminating from this treatment.

Our findings show a significant reduction in the level of IL-4 and TGF-β in co-infected (group 4), and helminth infected cattle (group 3) after deworming. Notably, our findings were corroborated by earlier studies carried out by Elias *et al*. 2001 and 2008 and others, where anthelminthic treatment was associated with improved BCG vaccination and reduced TGF-β production in helminth infected individuals (15, 17, 37). It has also been suggested that the removal of intestinal helminths by deworming would remove the inhibitory effects of Th2 on Th1 responses (38, 39). The capacity of helminth infection to modulate host’s immune system, particularly towards Th2 bias has been thoroughly elucidated by Maizel *et al*. 2004 (20). Previous experimental studies have reported an elevated IL-4 level, an important Th2 cytokine, in animals (5, 28) and humans (17) coinfected with helminth and mycobacteria. In general, the existing body of evidence indicates that helminths could bias immunity to mycobacteria infection (16, 55). This is supported by the association of elevated levels of Th2 cytokines, such as IL-4 and IL-13 (40, 41). Again, previous reports in human studies have indicated the association of helminths with enhanced regulatory T cell (Treg) activity characterised by increased expression of TGF-β (17). Importantly, several previous studies have implicated Tregs (TGF-β producing T cell) in the impairment of immune response during helminth concurrent infection with other pathogens (42, 43).

Drawing from our results, it can be deduced that prior screening for bTB among cattle in the study area, given the co-existence of helminths, would have culminated in the under-diagnosis of bTB using SICCTT. We show that application of anthelmintic treatment has the potential to reverse this down-modulation of BTB responses by helminths and allow for a more sensitive detection of the true rate of BTB infection.

Infected, undetected animals have the potential to spread the infection to other cattle within the herd, wildlife in the area and, if moved, to other herds in different parts of the country. Ultimately, this may pose a serious threat to public health through the zoonotic transmission of *M. bovis* to the unsuspecting public and various livestock workers and further frustrate the global effort of eradicating TB in humans. This becomes pertinent because the control, eradication and effective and/or successful treatment of any infectious disease rely solely on its accurate diagnosis.

## CONCLUSION

Our study showed that anti-helminthic treatment significantly improved the response to SICCTT in bTB-infected cattle. This response is characterized by an increase in IFN-γ production and reduction of IL-4 and TGF-β. The findings suggest that helminth infections may reduce the accuracy of current bTB diagnostic methods in cattle. The use of anthelmintics prior to SICCTT therefore might be a useful addition to bTB test and control programs. Finally, we suggest that further studies should be carried out to evaluate the anthelmintic treatment of cattle under different natural field conditions. These will help explore other nuances that may be associated with helminth-bTB co-infections that this study did not consider especially considering the relative few sample size used in this study.

## Data Availability

All the data used for this study will be made available on request by AND

## Acknowledgement

The authors wish to acknowledge the contributions of Dr. Shelley Rhodes for review of the manuscript and Prof. Martin Vordermeier for providing the PPDs used in carrying out this study. Both are of the Animal and Plant Health Agencies, United Kingdom.

## Authors Contributions

SC conceived the idea of the project, provided the reagents and materials used carried out critical review and approved the final draft of the manuscript. VOA carried out the field and laboratory work as well as the data analysis and wrote the first draft of the manuscript. TAJ, OT and OOA carried out the pathology. All authors read and approved the final version of the manuscript.

